# Anti-SARS-CoV-2 seropositivity among medical students in Copenhagen

**DOI:** 10.1101/2021.02.09.21251421

**Authors:** Johannes R. Madsen, Jacob P. S. Nielsen, Kamille Fogh, Cecilie B. Hansen, Pernille B. Nielsen, Theis Lange, Rasmus B. Hasselbalch, Peter Garred, Kasper Iversen

## Abstract

**Background:** Healthcare workers are at a higher risk of getting infected with SARS-CoV-2 than the general population. However, detailed knowledge about medical students and exposure to SARS-CoV-2 is lacking. Thus, we measured the prevalence of SARS-CoV-2 antibodies in a cohort of Danish medical students.

**Methods:** We invited all medical students at the University of Copenhagen (UCPH) to participate. A venous blood sample was drawn along with completing a self-report questionnaire. Blood samples were analyzed for total antibodies to SARS-CoV-2 and related to symptomatology, social behaviour, and work-life. Seropositive samples were screened for IgM, IgG, and IgA antibodies.

**Results:** Between October 19^th^ and 26^th^ 1,120 students participated in questionnaire, of these 1,096 were included in the study. Of all included 379 (34.58%) of the participants were seropositive. The risk of seropositivity was significantly increased for participants attending two parties at UCPH, on February the 29th and March 6^th^ 2020 (OR 5.96 (95% CI 4.34-8.24, p<0.001). Further, 461 students (42.06%) worked with COVID-19 patients, which was significantly associated with risk of seropositivity (OR 1.38, 95% CI 1.03-1.85, p=0.033). The symptom most substantially associated with seropositivity was loss of smell and/or taste (n=183 of all, 31.35%) with an OR of 24.48 (95% CI 15.49-40.60, p<0.001).

**Conclusion:** Medical students have the highest reported seropositivity in the Danish healthcare system. The risk of SARS-CoV-2 transmission amongst the students appear to be partly related to working with COVID-19 infected patients, but to a greater extent, their social behaviour.

**Mainpoints:** Medical students have the highest reported seropositivity in the Danish healthcare system. The risk of SARS-CoV-2 transmission amongst the students appear to be partly related to working with COVID-19 infected patients, but to a greater extent, their social behaviour.

## Introduction

Corona Virus Disease 2019 (COVID-19) caused by Severe Acute Respiratory Syndrome Corona Virus 2 (SARS-CoV-2), was on March 11^th^ 2020 declared a pandemic by WHO^1^, and by February 5^th^ 2021 more than onehundred and three millions have been infected and around 2.2 million have died because of COVID-19^2^.

In Denmark, medical students are employed as healthcare workers in hospitals during their studies and especially during the pandemic ^3^. Medical students assist the hospital departments in various functions such as substitute healthcare workers in clinical departments, as viral testers (nasopharyngeal and throat swabs), and aid in collecting blood samples for seroepidemiological studies ^4^. Furthermore, a large part of the Master’s programme at the University of Copenhagen (UCPH) is also based on clinical stays at hospitals in the Capital Region of Denmark^5^. Consequently, medical students encounter asymptomatic individuals infected with SARS-CoV-2 as well as Covid-19 patients.

An early study showed that the prevalence of antibodies among Danish medical students employed directly by the hospitals were a high-risk group with a SARS-CoV-2 seroprevalence of 14.97% ^6^.

However, since most medical students are not employed at hospitals, the overall seroprevalence of medical students is unknown. This also excludes them from the regional guidelines for systematic swap testing for SARS-CoV-2 of healthcare workers ^7^. Therefore, a potential unreported number of SARS-CoV-2 outbreaks amongst medical students is possible.

Additionally, medical students are in the age group with the highest incidence of positive SARS-CoV-2 PCR-test in Denmark ^8^. At March 6^th^ 2020 there was a party at the Faculty of Health and Medical Sciences at UCPH, where it was later confirmed that at least one SARS-CoV-2 infected medical student had attended ^9^. This also raises the possibility of the disease spreading rapidly and maybe even give rise to a superspreader event ^10^.

We aimed to examine the prevalence of antibodies against SARS-CoV-2 in medical students at UCPH and investigate the seroprevalence concerning their work and social life.

## Methods

### Participants

All medical students who were enrolled at UCPH (n=3673)^11^, were offered the possibility to participate in this observational cohort study. Medical students were invited by e-mail via their university e-mail sent by the union of Danish medical students (FADL).

Blood sampling took place between the 19^th^ and 26^th^ of October at 5 test locations. In relation to the sampling, the participants were asked to complete a questionnaire regarding symptomatology, information about demographics, type of work, history of contact with COVID-19 infected patients, as well as social life and general habits during the pandemic. The questionnaire was made in Research Electronic Data Capture (RedCap), and all the data collected was also managed by REDCap. REDCap is a secure, web-based tool hosted on servers owned by the Capital Region ^12, 13^.

### Blood samples

A venous blood sample of 6 ml was drawn from the antecubital region from all participants. Serum was stored at –80 °C until analysis and only thawed once.

A commercial ELISA method (Beijing Wantai Biological Pharmacy Enterprise, China) was used for determining seropositivity by measuring total antibodies to SARS-CoV-2 as previously described ^14 15^. The assay has a sensitivity of around 97% with a specificity of 99.5% in COVID-19 convalescent individuals. The samples that tested positive for total antibodies to SARS-CoV-2 went on to be analyzed using a direct ELISA method to detect IgG, IgA and IgM levels as described ^16^. The IgG, IgA and IGM direct ELISAs perform with sensitivities and specificities of 94.3 and 99.5%, 63.4 and 99.3%, and 61.4% and 99.1%, respectively. The numbers are obtained within 11 weeks of disease onset^16^.

### Ethics

The study complied with the Declaration of Helsinki II and was approved by the regional ethics committee of the Capital Region (H-20055767) and by the Danish Data Protection Authorities (P-2020-92). Following the guidelines for the provision of oral and written information, all patients gave written informed consent to participate in the study.

## Statistics

The primary outcome was the proportion of the study population with a positive antibody test for SARS-CoV-2. Calculations were done using R version 3.6.1 (R Foundation for Statistical Computing, Vienna, Austria 2019). Baseline characteristics and exposures were presented as n (%) for factors and mean (standard deviation (SD)) or median (interquartile range (IQR)) for numeric variables as appropriate.

To account for the expected clustering effect of medical students socialising within their semester, potential exposures variables (e.g. working with COVID-19 patients) were explored using multiple logistic regression models adjusted for semester of medical school and baseline characteristics significantly associated with seropositivity (age, sex and body mass index (BMI)). Results of these regressions were reported as odds ratios (OR) of seropositivity and presented with 95% confidence intervals (95% CI). A p-value of <0.05 was considered significant.

Similarly, we tested the association between symptoms and seropositity in univariate logistic regression models. Further, as COVID-19-like symptoms were expected to be common (e.g. runny nose) in the background population, we wished to estimate the prevalence of patients with SARS-CoV-2 antibodies without symptoms attributable to COVID-19. We calculated the probability of having symptoms of COVID-19 adjusted for the probability of seronegative participants having symptoms. This was done as previously described ^17^and details can be seen in the supplementary materials (See supplementary materials page 18-19).

## Results

From the 19th of October to the 26th of October a total of 1,120 medical students filled out the questionnaire and of these 1,096 had full serology performed and were included in the study, Figure 1 shows a CONSORT diagram of the study population. The mean age was 24.02 years (SD 3.52) and 782 (71.35%) were female. Of the included medical students 379 (34.58%, 95% CI 31.82-37.45) had anti SARS-CoV-2 antibodies. Among the seropositive 288 (76.19%, 95% CI 71.65-80.21) had detectable IgG antibodies, 10 (2.65%, 95% CI 1.44-4.8) had IgM antibodies and 58 (15.34%, 95% CI 12.06-19.32) had IgA antibodies. Table 1 shows baseline characteristics of the cohort divided according to seropositivity. The seropositive participants were significantly younger (23.48 vs 24.3, p<0.001), more likely to be male (41.08% for men vs 31.97% for women, p=0.01) and had a significantly lower BMI (22.67 vs 22.23, p=0.014). Figure 2 shows the percentage of seropostive for each semester of the medical school. Bachelor students (1st-6th semester students) were significantly more likely to be seropositive (42.28% vs 16.87%, p<0.001).

**Table 1.** Baseline characteristics

|  | Seronegative | Seropositive | p |
| --- | --- | --- | --- |
| **n** | **717** | **379** |  |
| **Age (mean (SD))** | 24.30 (3.95) | 23.48 (2.42) | <0.001 |
| **Female (%)** | 532 (74.2) | 250 (66.0) | 0.005 |
| **Body mass index (mean (SD))** | 22.23 (2.63) | 22.67 (2.90) | 0.011 |
| **Tobacco use (%)** | 64 ( 8.9) | 38 (10.0) | 0.585 |
| **Asthma (%)** | 76 (10.6) | 38 (10.0) | 0.835 |
| **Diabetes (%)** | 8 ( 1.1) | 3 ( 0.8) | 0.757 |
| **Other chronic lung disease (%)** | 2 ( 0.3) | 1 ( 0.3) | 1.000 |
| **Heart disease (%)** | 3 ( 0.4) | 1 ( 0.3) | 1.000 |
| **Kidney disease (%)** | 0 ( 0.0) | 2 ( 0.5) | 0.119 |
| **Weakened Immune system (%)** | 5 ( 0.7) | 5 ( 1.3) | 0.328 |
| **Hypertension (%)** | 5 ( 0.7) | 0 ( 0.0) | 0.171 |
| **Allergies (%)** | 224 (31.2) | 96 (25.3) | 0.043 |

*(Seropositive defined using the Wantai total Ig assay, Tobacco use defined as active smoker or user of snuff)*

**Figure 1.**
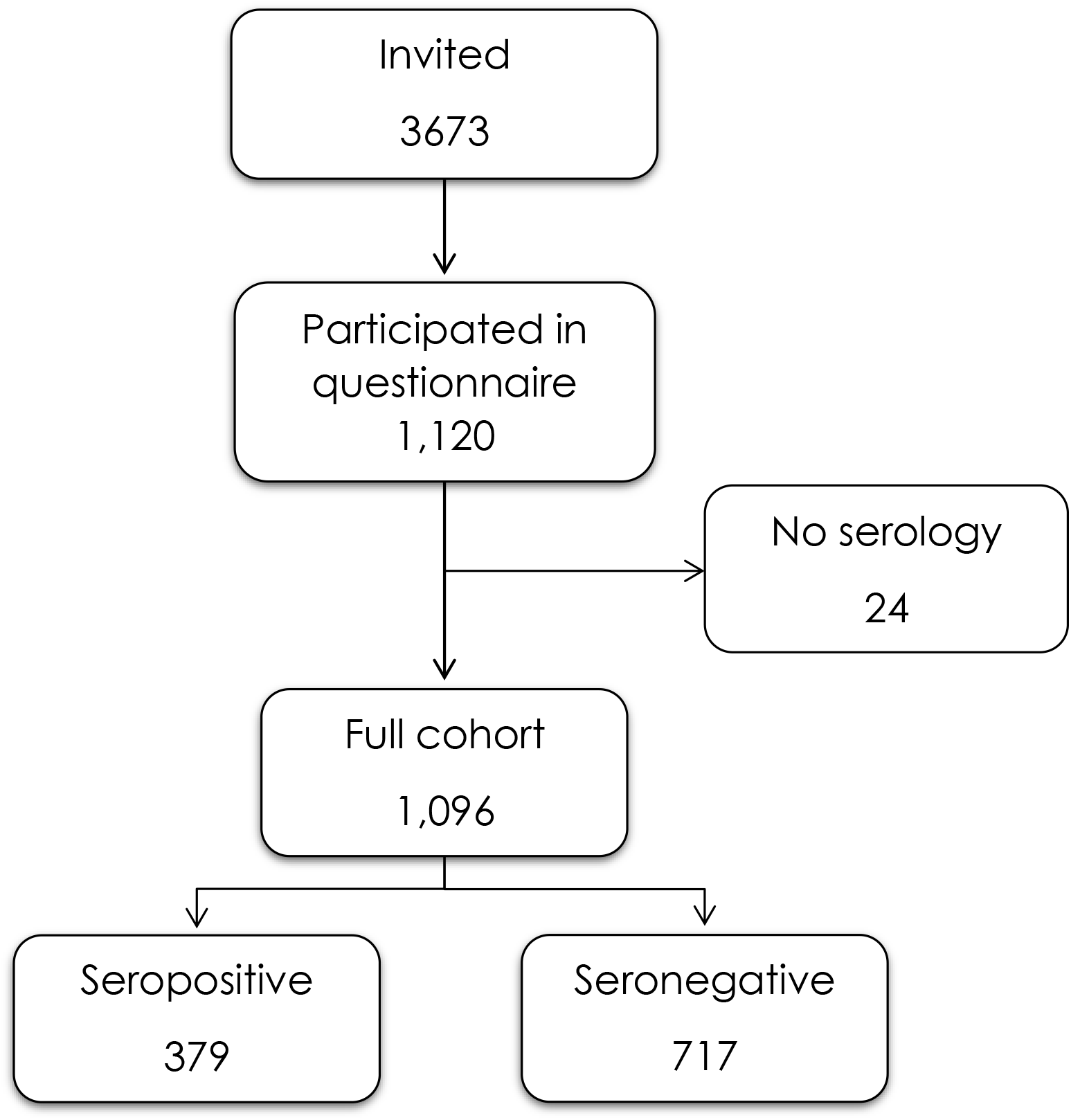
CONSORT diagram

**Figure 2.**
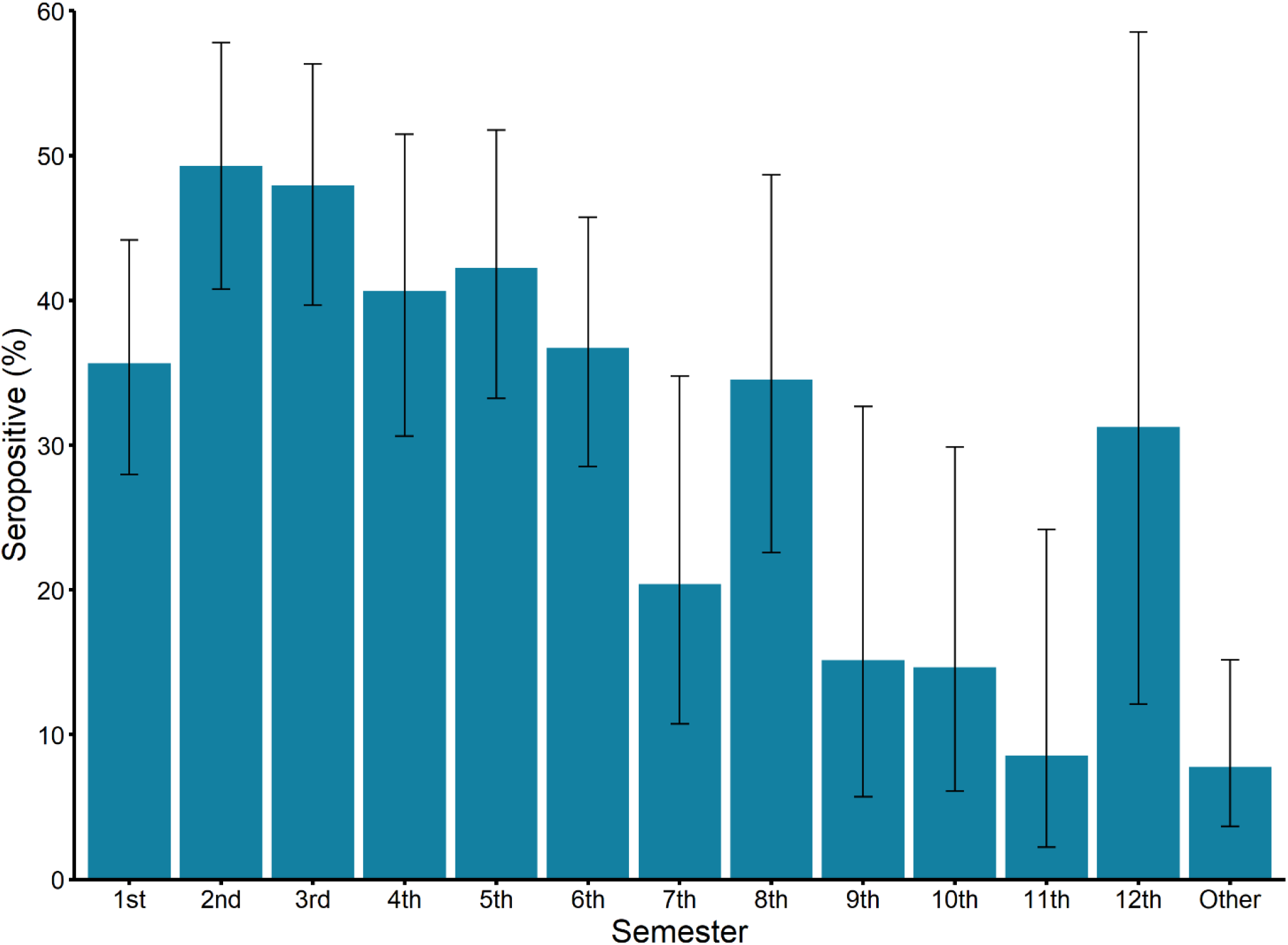
Risk of seropositivity according to semester. Errorbars indicate the 95% confidence interval

### Exposures

Table 2 shows potential exposures divided according to seropositivity. To correct for the expected clustering effect associated with socialising with other medical students at the same semester, a multivariate logistic model was used to determine the significance of the exposures. The results of this logistic regression is shown in table 3. Overall the risk of seropositivity was greatly increased for participants of the two parties held on the 29th of February 2020 and 6th of March 2020 at the Faculty of Health and Medical Sciences, UCPH. At the party on the 29th of February 272 (24.84%) of the cohort participated and of these 161 (59.19%) were seropositive resulting in an OR of 2.48 (95% CI 1.76-3.49, p<0.001). Similarly, for the party on the 6th of March 290 (26.48%) of the cohort participated and of these 181 (62.41%) were seropositive resulting in an OR of 3.69 (95% CI 2.68-5.1, p<0.001). Participating at either party was associated with an OR of 5.96 (95% CI 4.34-8.24, p<0.001) while participating in both parties was associated with an OR of 6.62 (95% CI 4.23-10.49, p<0.001). Working with COVID-19 patients was common in the cohort (n=461, 42.06%) and was associated with a significantly increased risk of seropositivity, with medical students who reported working with COVID-19 patients having a OR of 1.38 (95% CI 1.03-1.85, p=0.033). Of the baseline variables significantly associated with seropositivity BMI was the only one who remained significantly associated with seropositivity in the adjusted model with an OR of 1.07 (95% CI 1.02-1.13, p=0.012). Age and sex were not significant predictors of seropositivity.

**Table 2.** Exposures and risk

|  | Seronegative | Seropositive |
| --- | --- | --- |
| **n** | **717** | **379** |
| **Participated in party 6/3 (%)** | **109 (15.2)** | **181 (47.9)** |
| **Participated in party 29/2 (%)** | **111 (15.5)** | **161 (42.6)** |
| **Participated in any of the two parties (%)** | **170 (23.7)** | **257 (68.0)** |
| **Hours at institute (median [IQR])** | **8 [0, 25]** | **24 [6, 30]** |
| **Any work (%)** | **588 (82.0)** | **335 (88.4)** |
| **Any work with COVID-19 patients (%)** | **269 (37.5)** | **192 (50.7)** |
| **Work swabbing for COVID-19 (%)** | **48 (49.0)** | **42 (57.5)** |
| **Number of contacts a day (%)** |  |  |
| **1** | **199 (27.8)** | **119 (31.5)** |
| **2** | **276 (38.5)** | **142 (37.6)** |
| **3** | **146 (20.4)** | **60 (15.9)** |
| **4** | **52 ( 7.3)** | **27 ( 7.1)** |
| **5** | **24 ( 3.3)** | **13 ( 3.4)** |
| **6** | **20 ( 2.8)** | **17 ( 4.5)** |
| **More than 10 contacts a day (%)** | **96 (13.4)** | **57 (15.1)** |
| **Travel to area of risk (%)** | **67 ( 9.3)** | **44 (11.6)** |
| **Austria (%)** | **14 ( 2.0)** | **7 ( 1.8)** |
| **Italy (%)** | 12 ( 1.7) | 4 ( 1.1) |
| **France (%)** | 45 ( 6.3) | 35 ( 9.2) |

*(Seropositive defined using the Wantai total Ig assay, Travel risk areas were defined as travel to Italy, France, Austria or China during the period 1st of January to 1st of April, there were no travels to China)*

**Table 3.** Results of a multivariate logistic model of exposures and baseline characteristics

|  | OR | (95% CI) | p |
| --- | --- | --- | --- |
| **Age** | 0.99 | (0.93-1.04) | 0.671 |
| **Female** | 0.86 | (0.62-1.20) | 0.378 |
| **Body mass index** | 1.07 | (1.02-1.13) | 0.012 |
| **Working with COVID-19** | 1.38 | (1.03-1.85) | 0.033 |
| **Participated in party 29/2** | 2.48 | (1.76-3.49) | <0.001 |
| **Participated in party 6/3** | 3.69 | (2.68-5.10) | <0.001 |
| **Hours at institute (0-2 hours),**<br>**ref** | 1 |  |  |
| **Hours at institute (2-25**<br>**hours)** | 1.16 | (0.76-1.79) | 0.495 |
| **Hours at institute (>25 hours)** | 1.44 | (0.90-2.31) | 0.133 |

*(In addition to the abovementioned variables, the model was adjusted for the semester of the student (data not shown))*

Number of hours spent at the institute (the Faculty of Health and Medical Sciences, UCPH) or number of social contacts a day were not significant predictors of seropositivity. Likewise there was no increased risk for medical students who had traveled to either of the countries where the SARS-CoV-2 virus was most prevalent in the early spring, (France, Italy, Austria and China). However, there were relatively few travels to these countries. No medical students traveled to China during the study period.

### Symptoms

Having any illness during the period was significantly associated with seropositivity (601 of all, 54.39%) with an OR of 4.80 (95% CI 3.62-6.4, p<0.001). Still, 22.28% of seropositive participants reported no illness during the period. The symptom strongest associated with seropositivity was loss of smell and/or taste (n=183 of all, 31.35%) with an OR of 24.48 (95% CI 15.49-40.60, p<0.001). Table 4 shows the risk of seropositivity for participants who reported any illness, fever, loss of smell and/or taste and at least 3 symptoms. All reported symptoms and associated risk are shown in supplementary table 1.

**Table 4.** Selected symptoms and seropositivity

|  | All | (%) | Seropositive | (%) | OR | (95% CI) | p |
| --- | --- | --- | --- | --- | --- | --- | --- |
| **Any illness** | 596 | 54.48 | 294 | 77.78 | 4.8 | (3.62-6.4) | <0.001 |
| **Fever** | 345 | 31.48 | 193 | 50.92 | 3.86 | (2.95-5.06) | <0.001 |
| **Loss of taste/smell** | 182 | 16.61 | 161 | 42.48 | 24.48 | (15.49-40.6) | <0.001 |
| **At least 3 symptoms** | 526 | 47.99 | 264 | 69.66 | 3.99 | (3.06-5.22) | <0.001 |

When adjusting for the prevalence of symptoms among the seronegative participants 61.7% of seropositive participants had symptoms attributable to their infection.

## Discussion

Our results demonstrate that medical students at UCPH have a significantly higher prevalence of antibodies against SARS-CoV-2 in October 2020 than the general Danish population. In August 2020 the seroprevalence of SARS-CoV-2 antibodies in Denmark was 4.6% in the age group 20-29 years old^18^. A review from November 2020 showed an overall average seroprevalence of SARS-CoV-2 antibodies among healthcare workers on a global scale at 8.7%^19^. However, it is difficult to directly compare medical students and other healthcare workers both because of the possible difference in social behaviours and difference in working environments. Danish medical students often shift work enviroment between hospitals, wards, ect. We found that the risk of seropositivity was greatly increased for participants of two parties held on the 29th of February 2020 and the 6th of March 2020 at the Faculty of Health and Medical Sciences, UCPH, before the lockdown of Denmark on the 11th of March. This could potentially have been a superspreader event as well as the ones seen in USA^20^, Thailand ^21^ etc. These superspreader events can be very problematic, especially as 1/5 of the participants at the social gatherings for medical students did not have any disease symptoms. Therefore, these students were able to work at various hospitals all over the Capital Region of Denmark.

From the 1^st^ semester, medical students can work in the Danish healthcare system in various functions. This mainly happens by the union of Danish medical students’ employment agency (FADL’s vagtbureau). However, they are not employed at a specific department at a particular hospital, but instead, they work at various departments at different hospitals all over the Capital Region of Denmark. This gives great flexibility and creates a significant labour force for the hospitals. However, it also creates a risk as the medical students moves around.

Loss of taste and/or smell was a common symptom of COVID-19 infection in our study and it was highly associated with seropositivity with an OR of 24.48 (95% CI 15.49-40.60, p<0.001). Unlike other similar studies, the percentage of medical students who lost their taste and/or smell was considerably higher than others ^22-24^ (20.9 % vs. 16 % vs. 15 %). A considerable number of medical students with antibodies did not exhibit any symptoms, which raises the risk of asymptomatic disease transmission.

Very few of the medical students had detectable IgM and IgA antibodies (less than 3% and 16% of the seropositive, respectively). Since antigen specific IgM antibodies only appears in serum after the initial phase of infections we interpret this as an indication that most seropositive medical students had encountered the virus during the pandemic’s first wave in the spring 2020. Likewise, IgA antibodies in serum have been shown to decrease rapidly roughly 1 month after onset of COVID-19 symptoms. IgA antibodies are in general regarded as crucial in mucosal immunity and have furthermore been shown to be more abundant in saliva than in serum from COVID-19 infected individuals^25^. Among those positive, in the total Ig ELISA, around 76% were positive for IgG. We ascribe this discrepancy from the fact that the total Ig sandwich ELISA format provided by the Wantai system seems to be more sensitive than the direct IgG ELISA format as has been described previously^26^.

Strengths of this study include high participation (approximately 1/4 of the medical students at UCPH) and a high percentage of participants completed the questionnaire. All medical students were invited to participate regardless of previous antibody status or presence of symptoms. The study is limited by relying on participants’ retrospectively self-reported symptoms, social behavior etc., which allows for an unknown amount of misclassification.

In conclusion, we found that medical students at UCPH have an overall high seroprevalence of SARS-CoV-2 antibodies compared with other groups of healthcare workers and the general Danish population^6 18 27^. The risk of infection amongst the medical students appear to be partly related to working with COVID-19 infected patients, but to a greater extent, their social behaviour. This study points to the seriousness of superspreading events, especially when a large proportion of themedical students show no symptoms related to COVID-19 and therefore pose a risk of being asymptomatic carriers of the infection.

## Supporting information

Supplemental statistics + table

## Data Availability

No data are available

## Authors’ contributions

Conceived and designed the study; JRM, JPSN, KF, CBH, PBN, TL, RBH organized sample collection and analyzed the data; JRM, JPSN, PG and KI wrote the paper with inputs from all co-authors. All authors approved the final version of the manuscript.

## Acknowledgement

The authors would like to thank Camilla Xenia Holtermann Jahn, Sif Kaas Nielsen, and Jytte Bryde Clausen, from the Laboratory of Molecular Medicine at Rigshospitalet, for their excellent technical assistance. The authors want to thank all the volunteer medical students from UCPH that participated in the study.

## References

1. WHO. Rolling updates on coronavirus disease (COVID-19). Updated 31-07-2020. Accessed 02-12-2020, 2020. https://www.who.int/emergencies/diseases/novel-coronavirus-2019/events-as-they-happen

2. WHO. WHO Coronavirus Disease (COVID-19) Dashboard. Updated 04-02-2020. Accessed 05-02, 2020. https://covid19.who.int/

3. Rasmussen S, Sperling P, Poulsen MS, Emmersen J, Andersen S. Medical students for health-care staff shortages during the COVID-19 pandemic. Lancet. May 2 2020;395(10234):e79–e80. doi:10.1016/s0140-6736(20)30923-5

4. vagtbureau Fs. Om Vagtbureauet. 20–12, 2020. https://fadlvagt.dk/om-vagtbureauet/

5. Copenhagen Uo. Undervisning og opbygning. Accessed 21-12, 2020. https://studier.ku.dk/kandidat/medicin/undervisning-og-opbygning/

6. Iversen K, Bundgaard H, Hasselbalch RB, et al. Risk of COVID-19 in health-care workers in Denmark: an observational cohort study. Lancet Infect Dis. Dec 2020;20(12):1401–1408. doi:10.1016/S1473-3099(20)30589-2

7. Rigshospitalet. Systematisk COVID-19-test af ansatte med patientkontakt. Accessed 02-12, 2020. https://www.rigshospitalet.dk/for-fagfolk/corona/Sider/Systematisk-corona-test-af-ansatte-med-patientkontakt.aspx

8. Institut SS. Statens Serum Institut - covid-19 - Danmark. Accessed 02-12-2020, 2020. https://experience.arcgis.com/experience/aa41b29149f24e20a4007a0c4e13db1d

9. Authority DPS. COVID-19-smittet var til fredagsbar på Panum Instituttet i København fredag 6. marts. Danish Patient Safety Authority. Updated 10-03-2020. Accessed 02-12-2020, 2020. https://stps.dk/da/nyheder/2020/covid-19-smittet-var-til-fredagsbar-paa-panum-instituttet-i-koebenhavn-fredag-6-marts/?fbclid=IwAR0FFElUCtLHSMtPh3jg-mOWN-yO7HQciv_hg8Z5GH1DrIR2Hb5L7CMLAYw#

10. Kochańczyk M, Grabowski F, Lipniacki T. Super-spreading events initiated the exponential growth phase of COVID-19 with ℛ(0) higher than initially estimated. R Soc Open Sci. Sep 2020;7(9):200786. doi:10.1098/rsos.200786

11. Copenhagen Uo. Bestand. 13-01, 2021. Updated 01-10-2020. Accessed 13-01, 2021. https://us.ku.dk/studiestatistik/studiestatistikker/bestand/?fbclid=IwAR11VkxjefOzUFQhrekNzrms2U5KBsN7TM4PJdHrCO_CP39Y4z_BJsOa4lY

12. Harris PA, Taylor R, Minor BL, et al. The REDCap consortium: Building an international community of software platform partners. Journal of Biomedical Informatics. 2019/07/01/ 2019;95:103208. doi:https://doi.org/10.1016/j.jbi.2019.103208

13. Harris PA, Taylor R, Thielke R, Payne J, Gonzalez N, Conde JG. Research electronic data capture (REDCap)—A metadata-driven methodology and workflow process for providing translational research informatics support. Journal of Biomedical Informatics. 2009/04/01/ 2009;42(2):377–381. doi:https://doi.org/10.1016/j.jbi.2008.08.010

14. Harritshoej LH, Gybel-Brask M, Afzal S, et al. Comparison of sixteen serological SARS-CoV-2 immunoassays in sixteen clinical laboratories. medRxiv. 2020:2020.07.30.20165373. doi:10.1101/2020.07.30.20165373

15. Weidner L, Gansdorfer S, Unterweger S, et al. Quantification of SARS-CoV-2 antibodies with eight commercially available immunoassays. J Clin Virol. Aug 2020;129:104540. doi:10.1016/j.jcv.2020.104540

16. Hansen CB, Jarlhelt I, Perez-Alos L, et al. SARS-CoV-2 Antibody Responses Are Correlated to Disease Severity in COVID-19 Convalescent Individuals. J Immunol. Nov 18 2020;doi:10.4049/jimmunol.2000898

17. D’Souza M, Nielsen D, Svane IM, et al. The risk of cardiac events in patients receiving immune checkpoint inhibitors: a nationwide Danish study. European heart journal. Dec 9 2020;doi:10.1093/eurheartj/ehaa884

18. Institut SS. Covid-19: Den nationale Prævalensundersøgelse. 07-10-2020 2020;

19. Galanis P, Vraka I, Fragkou D, Bilali A, Kaitelidou D. Seroprevalence of SARS-CoV-2 antibodies and associated factors in health care workers: a systematic review and meta-analysis. J Hosp Infect. Nov 16 2020;doi:10.1016/j.jhin.2020.11.008

20. Lemieux JE, Siddle KJ, Shaw BM, et al. Phylogenetic analysis of SARS-CoV-2 in Boston highlights the impact of superspreading events. Science. Dec 10 2020;doi:10.1126/science.abe3261

21. Chau NVV, Hong NTT, Ngoc NM, et al. Superspreading Event of SARS-CoV-2 Infection at a Bar, Ho Chi Minh City, Vietnam. Emerg Infect Dis. Jan 2021;27(1)doi:10.3201/eid2701.203480

22. Foster KJ, Jauregui E, Tajudeen B, Bishehsari F, Mahdavinia M. Smell loss is a prognostic factor for lower severity of coronavirus disease 2019. Ann Allergy Asthma Immunol. Oct 2020;125(4):481–483. doi:10.1016/j.anai.2020.07.023

23. Lan FY, Filler R, Mathew S, et al. COVID-19 symptoms predictive of healthcare workers’ SARS-CoV-2 PCR results. PLoS One. 2020;15(6):e0235460. doi:10.1371/journal.pone.0235460

24. Lee Y, Min P, Lee S, Kim SW. Prevalence and Duration of Acute Loss of Smell or Taste in COVID-19 Patients. J Korean Med Sci. May 11 2020;35(18):e174. doi:10.3346/jkms.2020.35.e174

25. Sterlin D, Mathian A, Miyara M, et al. IgA dominates the early neutralizing antibody response to SARS-CoV-2. Sci Transl Med. Dec 7 2020;doi:10.1126/scitranslmed.abd2223

26. Bal A, Pozzetto B, Trabaud MA, et al. Evaluation of high-throughput SARS-CoV-2 serological assays in a longitudinal cohort of patients with mild COVID-19: clinical sensitivity, specificity and association with virus neutralization test. Clin Chem. Jan 5 2021;doi:10.1093/clinchem/hvaa336

27. Institut SS. Næsten en halv million danskere deltog i “Vi Tester Danmark”. Nu går fase 2 af projektet i gang. Updated 16. december 2020. Accessed 21-12, 2020. https://www.ssi.dk/aktuelt/nyheder/2020/nasten-en-halv-million-danskere-deltog-i-vi-tester-danmark-nu-gar-fase-2-af-projektet-gar-i-gang

