## Supplemental statistics + table for "Anti-SARS-CoV-2 seropositivity among medical students in Copenhagen"

### Supplementary materials

#### Supplementary statistics

Adjusting for symptoms among participants without COVID-19

We wished to estimate the prevalence of participants with SARS-CoV-2 antibodies without symptoms attributable to COVID-19. As symptoms of COVID-19 were expected to be common (e.g. runny/stuffed nose) in the background population, we chose to adjust the proportion of seropositive participants with symptoms to the proportion of seronegative patients with symptoms.

To calculate this, we had to make some assumptions/approximations:

1. We assumed perfect test characteristics (sensitivity and specificity) and thus no SARS-CoV-2 positive participants among those who tested negative by our test.
2. We assumed the risk of having COVID-19-like symptoms not attributable to COVID-19 to be equal among seropositive and seronegative participants
3. We assumed no recall bias regarding symptoms.

Given these assumptions we wanted to estimate the probability of having symptoms attributable to COVID-19 when seropositive:

$P(B)$

The probability of having COVID-19-like symptoms while seronegative is self-reported:

$$P(A)$$

The probability of being completely asymptomatic and seropositive is self-reported:

$$P(C)$$

For a participant to be completely asymptomatic and seropositive (P(C)), they would have to have no symptoms attributable to COVID-19 (1-P(B)) in addition to having no COVID-19-like symptoms during the period (1-P(B)). Given this, the probability of being completely asymptomatic and seropositive is equal to the probability of having no COVID-19-like symptoms while seronegative multiplied with the probability of not having symptoms attributed to COVID-19 and seropositive. Given this, the probability of being completely asymptomatic and seropositive is equal to the probability of having no COVID-19-like symptoms while seronegative multiplied with the probability of not having symptoms attributed to COVID-19 and seropositive:

$$P(C) = (1 - P(A)) \times (1 - P(B))$$

From here we can find the risk of symptoms as:

$$P(B) = 1 - \frac{P(C)}{1 - P(A)}$$

This way we estimate the risk of having symptoms attributable to COVID-19 adjusted for the risk of having similar symptoms while not infected.

#### Suppl table 1, other symptoms

| | All | (%) | Seropositive | (%) | OR | (95% CI) | p |
|---|---|---|---|---|---|---|---|
| **Chills** | 195 | 17.79 | 116 | 30.61 | 3.56 | (2.59-4.92) | <0.001 |
| **Runny/stuffed nose** | 326 | 29.74 | 149 | 39.31 | 1.98 | (1.51-2.58) | <0.001 |
| **Sneezing** | 205 | 18.70 | 82 | 21.64 | 1.33 | (0.97-1.82) | 0.071 |
| **Sore throat** | 407 | 37.14 | 172 | 45.38 | 1.7 | (1.32-2.20) | <0.001 |
| **Coughing** | 338 | 30.84 | 160 | 42.22 | 2.21 | (1.70-2.89) | <0.001 |
| **Shortness of breath** | 87 | 7.94 | 54 | 14.25 | 3.44 | (2.20-5.46) | <0.001 |
| **Headache** | 362 | 33.03 | 189 | 49.87 | 3.13 | (2.40-4.08) | <0.001 |
| **Myalgia and/or joint pain** | 221 | 20.16 | 137 | 36.15 | 4.27 | (3.14-5.83) | <0.001 |
| **Chest pain** | 40 | 3.65 | 29 | 7.65 | 5.32 | (2.7-11.24) | <0.001 |
| **Lethargy** | 384 | 35.04 | 218 | 57.52 | 4.49 | (3.44-5.88) | <0.001 |
| **Loss of appetite** | 130 | 11.86 | 79 | 20.84 | 3.44 | (2.37-5.04) | <0.001 |
| **Colored sputum** | 43 | 3.92 | 15 | 3.96 | 1.01 | (0.52-1.90) | 0.966 |
| **Conjunctivitis** | 21 | 1.92 | 12 | 3.17 | 2.57 | (1.08-6.35) | 0.034 |
| **Nausea** | 87 | 7.94 | 43 | 11.35 | 1.96 | (1.26-3.04) | 0.003 |
| **Vomiting** | 26 | 2.37 | 10 | 2.64 | 1.19 | (0.52-2.61) | 0.674 |
| **Diarrhea** | 59 | 5.38 | 34 | 8.97 | 2.73 | (1.61-4.69) | <0.001 |
| **Stomach pain** | 74 | 6.75 | 36 | 9.50 | 1.88 | (1.16-3.02) | 0.009 |

| | | | | | | | | |
|---|---|---|---|---|---|---|---|---|
| **Other** | 17 | 1.55 | | 9 | 2.37 | 2.16 | (0.82-5.79) | 0.117 |
